# Utilization of Triglyceride Levels and Estimated Average Plasma Glucose Using Glycated Hemoglobin in Assessing Insulin Resistance: TyHBA1c index

**DOI:** 10.1101/2023.12.20.23300240

**Authors:** Luís Jesuino de Oliveira Andrade, Luiz Felipe Moreno de Brito, Gabriela Correia Matos de Oliveira, Luisa Correia Matos de Oliveira, Alcina Maria Vinhaes Bittencourt, Gustavo Magno Baptista, Catharina Peixoto Silva, Luís Matos de Oliveira

**Affiliations:** Colegiado de Medicina - Departamento de Saúde Universidade Estadual de Santa Cruz - Ilhéus – Bahia – Brazil; Laboratório de Análises e Pesquisas – Itabuna – Bahia – Brazil; Programa Saúde da Família – Bahia – Brazil; Centro Universitário SENAI CIMATEC – Salvador – Bahia - Brazil; Faculdade de Medicina Universidade Federal da Bahia – Salvador – Bahia – Brazil; Clínica de Pneumologia – Itabuna – Bahia – Brazil; Escola Bahiana de Medicina e Saúde Pública - Salvador –Bahia –Brazil

**Keywords:** HOMA-IR, Triglyceride glucose index, HbA1c, Triglyceride HbA1c index

## Abstract

**Introduction:** Insulin resistance (IR) is a metabolic disorder characterized by reduced sensitivity to the physiological effects of insulin, leading to an impaired ability of cells to take up glucose from the bloodstream. There are multiple indices available for assessing IR, however, they often rely on a single glycemic parameter at a specific moment, leaving a gap in the evaluation of IR over a certain period.

**Objective:** To develop a novel index for assessing IR by combining triglyceride levels and estimated glycemia through glycated hemoglobin (HBA1c).

**Method:** We used the fasting triglycerides and glucose levels (TyG) index formula by replacing fasting glucose with estimated average glucose calculated through HbA1c. The study included 200 laboratory samples, including fasting glucose, triglycerides, insulin, homeostasis model assessment for IR (HOMA-IR) index, TyG index, and estimated average glucose by HbA1c. The ideal cut-off value of the TyHBA1c index for estimating IR was established compared to the HOMA-IR index, and TyG index using a receiver operating characteristic (ROC) analysis. A weighted kappa test was used to estimate the diagnostic agreement between the TyHBA1c index and the HOMA-IR and Tyg index.

**Results:** The TyHBA1c index showed a higher correlation with the HOMA-IR index (r = 0.273, p = 0.000) compared to the TyG index (r = -0.617, p < 0.000). An association between HBA1c and the HOMA-IR index was observed (r = 0.215, p = 0.002). The ROC curve showed that the cutoff value for the TyHBA1c index to best estimate IR is Ln 4.74 (sensitivity 85.0% and specificity 95.0%) compared to the TyG index. The weighted kappa test revealed moderate agreement between the TyHBA1c index and the HOMA-IR index (k = 0.45, p = 0.006), and satisfactory agreement between the TyHBA1c index and the TyG index (k = 0.71, p = 0.009).

**Conclusion:** Our results demonstrated a strong association between the TyHBA1c index and TyG index, and the HOMA-IR index, suggesting that the TyHBA1c index may be more a indicator of IR, and more a tool in clinical practice for assessing IR.

## Introduction

Insulin resistance (IR) is a metabolic disorder characterized by reduced sensitivity to the physiological effects of insulin, leading to an impaired ability of cells to take up glucose from the bloodstream.^1^ It is a key feature in the development of various diseases, including type 2 diabetes mellitus (T2DM), metabolic syndrome, and cardiovascular disorders.^2^ Accurate assessment of IR is crucial for early diagnosis and effective management of these conditions.

IR arises from the complex interplay between genetic and environmental factors. It is commonly associated with obesity, sedentary lifestyle, and poor dietary habits. The fundamental mechanism underlying IR involves the impaired signaling pathway of the insulin receptor, resulting in decreased glucose uptake and utilization by peripheral tissues such as skeletal muscle and adipose tissue.^3^

Triglycerides have been observed to have a significant impact on IR.^4^ Elevated levels of triglycerides contribute to the accumulation of fatty acids in non-adipose tissues, particularly in skeletal muscle and liver. This ectopic lipid deposition leads to the disruption of insulin signaling pathways and further exacerbates IR.^5^

The estimation of glycemia through glycated hemoglobin (HbA1c) provides valuable information about long-term glycemic control.^6^ HbA1c reflects the average blood glucose levels over the previous two to three months, making it an important marker for monitoring diabetes management.^7^ Studies have shown a strong association between elevated HbA1c levels and IR, suggesting that HbA1c could serve as a reliable surrogate for assessing IR.^8^

The euglycemic clamp technique is considered the gold standard for directly measuring insulin sensitivity.^9^ The homeostasis model assessment for IR (HOMA-IR) index is a widely used tool to assess IR in individuals.^10^ Both the HOMA-IR index and the euglycemic clamp play crucial roles in assessing IR and evaluating interventions to improve insulin sensitivity.

Recently, a new index of IR, called the Triglyceride-Glucose (TyG) index, has been proposed as a simple and reliable tool for the assessment of IR.^11^ Several studies have demonstrated a strong association between the TyG index and IR, as well as its ability to identify individuals at risk of developing diabetes.^12^

The TyG index takes into account the levels of both triglycerides and glucose, which are key components of IR. Elevated triglyceride levels indicate an abnormality in lipid metabolism, often seen in insulin-resistant individuals. Moreover, elevated glucose levels reflect impaired glucose metabolism, indicating the presence of IR. By combining these two parameters, the TyG index provides a comprehensive assessment of IR, making it a valuable tool for clinical practice and research.

The objective of this study is to develop a novel index of IR that combines the measurements of triglycerides and estimated glycemia through HbA1c (TyHBA1c index). By incorporating these two parameters, we aim to create a more comprehensive and accurate measure of IR. This index could potentially improve the diagnosis and management of IR-related conditions.

## Method

The HOMA-IR index was derived using the following formula: (Fasting insulin in betaU/mL × fasting glucose in mg/dL) / 415.^10^ In accordance with earlier research, an HOMA-IR index ≥ 3.4 indicates IR, which represents the optimal threshold for predicting the development of diabetes and aligns with the hyperglycemic-hyperinsulinemic clamp method. Furthermore, the TyG index was determined as Ln[fasting triglycerides (mg/dL) fasting glucose (mg/dL)/2], and it is expressed on a logarithmic scale.^11^ The benchmark for identifying IR is established at a TyG index value of 4.49.

The TyHBA1c index was calculated based in TyG index formula by replacing fasting glucose with estimated average glucose calculated through HbA1c.

### Sample Description

The study consisted of 200 samples, whose data were collected from laboratory records. Demographic data, including sex, age, fasting glucose, triglyceride levels, insulin levels, HOMA index, TyG index, HBA1C, and estimated mean glucose by HBA1c, were based on the availability of complete laboratory results.

### Statistical Analysis

Descriptive statistics were used to summarize demographic characteristics. Continuous variables were expressed as mean ± standard deviation depend. The categorical variable (gender) was presented as frequency and percentage.

The correlation between TyHBA1c index and HOMA-IR index was assessed using Pearson’s correlation coefficient.

The ideal cut-off value of the TyHBA1c index for estimating IR was established compared to the HOMA-IR index, and TyG index using a receiver operating characteristic (ROC) analysis. Sensitivity, specificity, true positive and negative predictive values, as well as positive and negative likelihood ratios of the TyHBA1c index were determined.

A weighted kappa test was used to estimate the diagnostic agreement between the TyHBA1c index and the HOMA-IR and TyG index. Statistical significance was set at p < 0.05.

### Ethical Considerations

The study was conducted according to the Declaration of Helsinki. The National Commission for Research Ethics (CONEP - Brazil) approved the project (registry number: 2.464.513). Confidentiality and data protection protocols were strictly followed to ensure patient privacy.

## Results

A total of 200 samples were evaluated, consisting of 126 (63.0%) females and 74 (37.0%) males, with a mean age of 46.56 ± 18.98 years. IR was identified in 64 (32.0%) using the HOMA-IR index and in 132 (66.0%) using the TyG index. The HOMA-IR and TyG indices did not exhibit similar behavior in the evaluated samples. The demographic characteristics are summarized in Table 1.

**Table 1.**

| **Gender** | **Male: 74 (37%) Female: 126 (63%)** |
| --- | --- |
| **Age (years)** | $46.56 \pm 18.98$ |
| **HBA1c** | $5.78 \pm 0.91$ |
| **Insulin** | $12.06 \pm 9.12$ |
| **Fasting glucose** | $106.90 \pm 23.24$ |
| **Average blood glucose estimated by HBA1C** | $119.26 \pm 25.99$ |
| **Triglycerides** | $124.84 \pm 66.63$ |
| **HOMA-IR** | $3.13 \pm 2.74$ |
| **TyG index** | $4.64 \pm 0.30$ |
| **TyHBA1c** | $4.74 \pm 0.30$ |

The association between the TyHBA1c index and the HOMA-IR index, as assessed by Pearson’s correlation (*r* = 0,273 and p = 0.000) was higher than correlation between TyHBA1c index and the TyG index (*r* = -,617 and p 0.000) (Table 2).

**Table 2.**
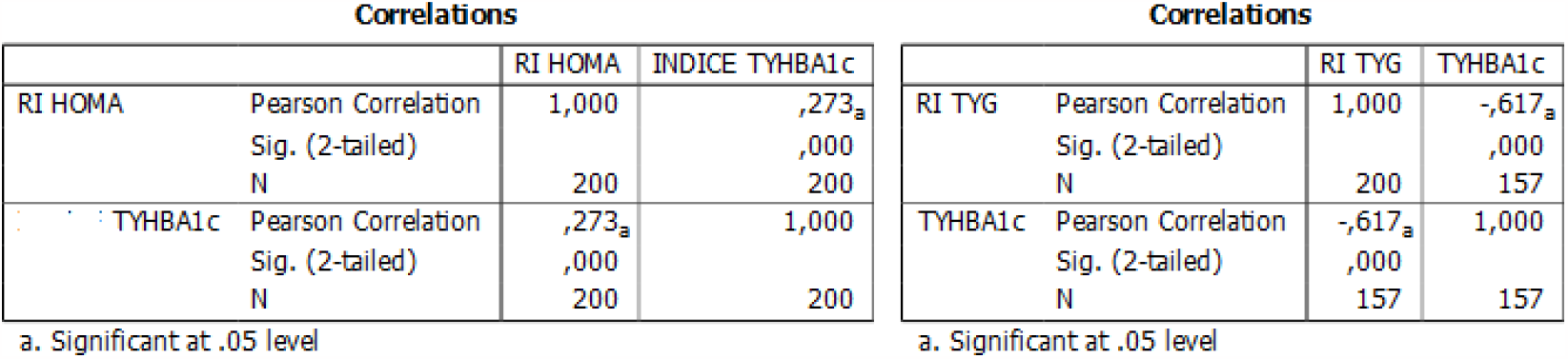
Correlation between the TyHBA1c index, HOMA-IR index and TyG index.

There was an association between the HBA1c and the HOMA-IR index, as assessed by Pearson’s correlation (r = 0,215 and p = 0.002).

ROC curve showed that the cut-off value best of the TyHBA1c index compared with the HOMA-IR index for estimating IR corresponds to Ln 4.57 (sensitivity 50.0% and specificity 23.0%) (Figure 1).

**Figure 1.**
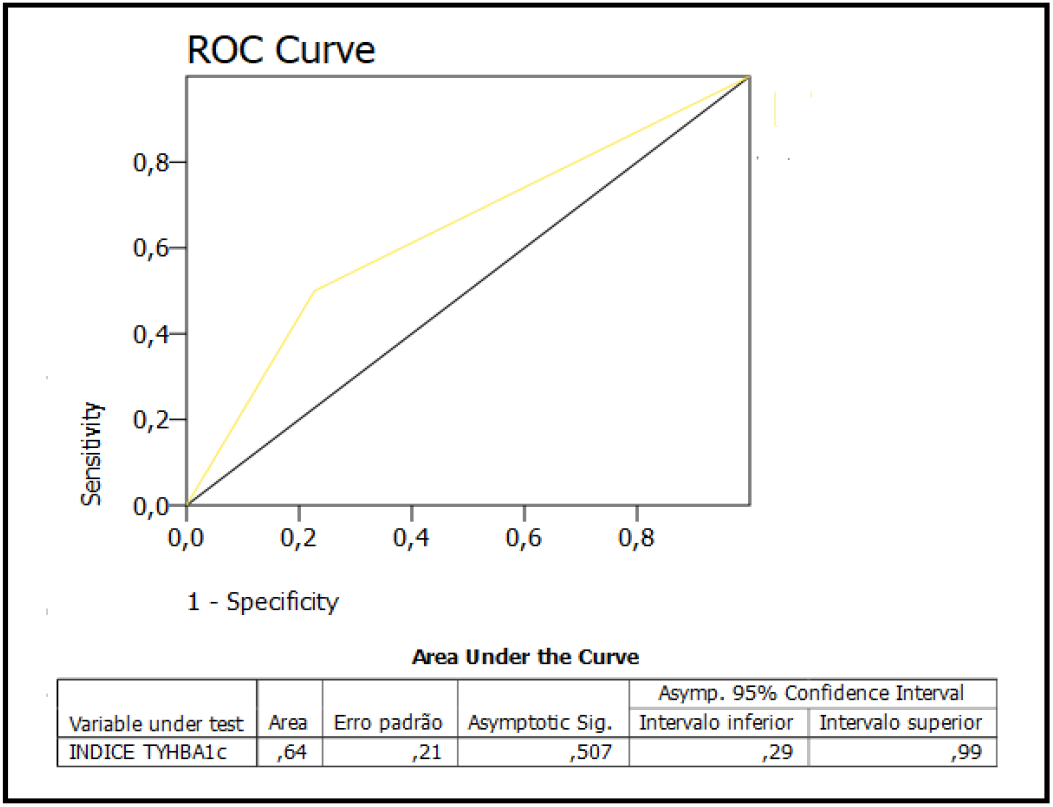
ROC analysis: TyHBA1c index compared to the HOMA-IR index

ROC curve showed that the cut-off value best of the TyHBA1c index compared with the TyG index for estimating IR corresponds to Ln 4.74 (sensitivity 85.0% and specificity 95.0%) (Figure 2).

**Figure 2.**
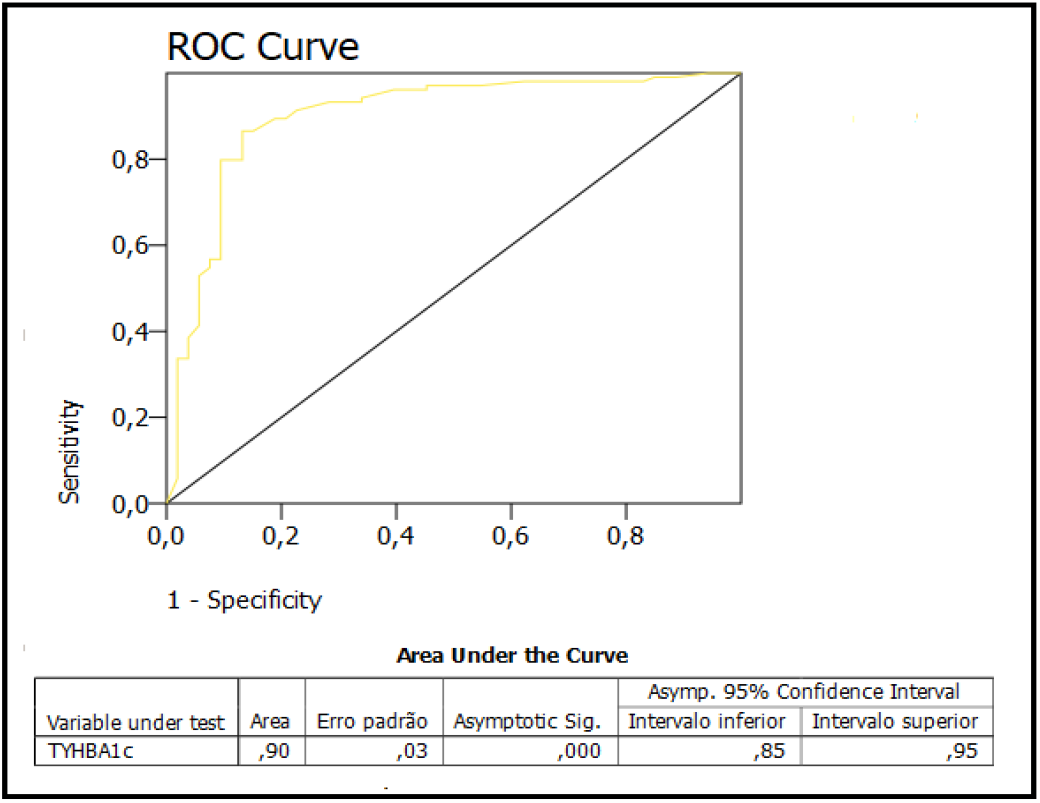
ROC analysis: TyHBA1c index compared to the TyG index

The weighted kappa test revealed moderate agreement between the TyHBA1c index and the HOMA-IR index (k = 0.45; p = 0,006), and demonstrated satisfactory agreement between the TyHBA1C index and the TyG index (k = 0.71; p = 0,009).

## Discussion

Our study evaluated whether the product of triglyceride levels and the estimated average glucose through HbA1c can be used as a substitute for the TyG index to assess insulin resistance, indicating that the HbA1c index demonstrates high sensitivity and specificity when compared to the TyG index. In a research study, sensitivity refers to the ability of a diagnostic test to accurately identify individuals with the condition of interest, while specificity pertains to its ability to accurately exclude individuals without the condition. Sensitivity represents the true positive rate, while specificity depicts the true negative rate. These metrics are crucial in evaluating the reliability and validity of diagnostic tests in clinical practice.^13^

The widely used HOMA-IR index has high sensitivity and specificity in estimating the risk of insulin resistance when compared to other more complex tests.^14^ In our study, the TyHBA1c index, when compared to the HOMA-IR index for estimating insulin resistance, showed moderate sensitivity and low specificity.

The HOMA-IR index and TyG index are commonly used to assess IR, with the former primarily focusing on the relationship between fasting glucose and insulin levels, while the latter incorporates triglyceride levels into its calculation.^10,11^ Our study has shown that using estimated average glucose derived from HbA1c in place of glucose levels in the TyG index can yield more accurate assessments of IR, as this approach offers higher sensitivity and specificity, thus improving the precision of IR evaluation, considering that HbA1c represents the average glucose levels over the past three months.

Studies have shown that increasing evidence indicates that the TyG index is correlated with HOMA-IR index and hyperinsulinemic-euglycemic clamp test.^15^ In our study, the association between the TyHBA1c index and HOMA-IR index, assessed by Pearson correlation, was superior to the correlation between the TyHBA1c index and the TyG index. However, weighted kappa test revealed moderate agreement between the TyHBA1c index and HOMA-IR index, and demonstrated satisfactory agreement between the TyHBA1c index and the TyG index.

The HbA1c is well established as a marker of glycemic control, and has the potential to reflect the history of average IR in the preceding weeks or months, whereas the TyG index and the HOMA-IR index reflect the current status of IR.^16^ In our study, the evaluation of the correlation between HbA1c and HOMA-IR index revealed a positive association between these variables. So we used the utilization of the estimated average glucose through HbA1c in the formula for the TyG index demonstrates a superior response in diagnosing IR compared to using current glucose levels.

The new insulin resistance marker index, TyHBA1C, demonstrated significant association when compared to the TyG index and the HOMA-IR index. This new marker may be a promising tool, as it utilizes an estimated average blood glucose level over the past three months, thereby offering greater precision in evaluating IR. Therefore, our results suggest that the TyHBA1C index may aid in early detection, monitoring, and management of conditions related to IR, potentially serving as an additional tool in clinical practice.

## Conclusion

Our results demonstrated a strong association between the TyHBA1c index and TyG index, and the HOMA-IR index, suggesting that the TyHBA1c index may be more a indicator of IR, and more a tool in clinical practice for assessing IR. However, further validation studies are needed to fully establish the clinical utility and potential integration of TyHBA1C into routine clinical practice.

## Data Availability

All data produced in the present work are contained in the manuscript

